# Questionnaire based Prediction of Hypertension using Machine Learning

**DOI:** 10.1101/2020.06.18.20133397

**Authors:** Abhijat Chaturvedi, Siddharth Srivastava, Astha Rai, A S Cheema, Desham Chelimela, Rajeev Aravindakshan

**Author notes:** {, }.

## Abstract

Machine Learning has proven its ability in healthcare as an assisting technology for health care providers either by saving precious time or timely alerts or vitals monitoring. However, their application in real world is limited by availability of data. In this paper, we show that simple machine learning algorithms especially neural networks, if designed carefully, are extremely effective even with limited amount of data. Specifically with exhaustive experiments on standard Modified National Institute of Standards and Technology database (MNIST) dataset we analyse the impact of various parameters for effective performance. Further, on a custom dataset collected at a tertiary care hospital for hypertension analysis, we apply these design considerations to achieve better performance as compared to competitive baselines. On a real world dataset of only a few hundred patients, we show the effectiveness of these design choices and report an accuracy of 75% in determining whether a patient suffers from hypertension.

## 1 Introduction

Hypertension is one of the most common ailment in the Indian patients and the severe threat that it poses makes it important to identify it as early as possible in order to stop it from becoming incurable. Fourth National Family Health Survey reports more than 13% of men and more than 8% of women in early to middle ages are suffering from hypertension [1]. The scarcity of healthcare experts makes the situation even more grim.

In today’s world machine learning becomes an obvious choice to solve such problems. It has already influenced every aspect of healthcare with major progress reported in for diseases prediction [2], cancer classification [3], drug discovery [4] etc. However with respect to the current work, these solutions have the following limitations (i) They work on extremely large data with approximately 40, 000 and above instances in the dataset [5] (ii) The data is mostly of non Indian patients. Studies indicate that there is disparity in healthcare with respect to race and ethnicity [6] and hence, it is yet to be established if the same set of techniques can generalize across them.

In this paper, we target the problem of prediction of hypertension. To diagnose hypertension in practice, a healthcare professional asks a series of questions to the patient and on the basis of answers it is identified if a patient is suffering from hypertension or not. We follow a similar approach, where a predefined set of questions were asked to several patients and their response was recorded. In view of this, following are the contributions of this paper.

- We evaluate the impact of hyperparameter choices on the performance of neural networks. With exhaustive experiment on MNIST, we evaluate them and provide insights on their impact.
- We collect a dataset for hypertension analysis and use the insights from the analysis to design a simple neural network providing better results than competitive baselines.

The rest of the paper is organised as follows. Section 2 contains Related Works, Section 3 discusses background of neural network, compared techniques and notations used in the paper. Section 4 contains designing of neural networks, Section 5 explains the experiments done with the models and dataset. Section 6 contains Results and Section 7 contains Conclusion.

## 2 Related Works

Machine learning algorithms are used in predicting diseases [7]. Diabetes prediction is an example of diseases prediction using machine learning [8]. Feras Hatib et. al. [9] used machine learning model for predicting hypertension using high-fidelity arterial waveforms. X Li et. al. [10] in their paper used feature extraction from sequential data and contextual data using hidden layers of recurrent structure and predicting the blood pressure value. They have used Root Mean Square Error as the evaluation metrics for their model. The image based prediction in healthcare machine learning is helping in predicting diseases like cancer [11][12]. This work is not intended in image based diseases classification, but for the sake of completeness and future progress in this work, image based studies are also referred.

Chengyin Ye1 et. al. [13] uses XGBoost for model creation in hypertension prediction. The dataset they used contains 1504437 instances from 2013 and 2014. However they classified the instances into 5 classes ranging from *Lowest risk* to *Very High Risk*. They have used Area Under Curve as the performance metrics. Mevlut Ture et. al. [14] presented a comparative study of machine learning algorithms for predicting hypertension. They have evaluated decision trees, statistical model and neural network on a dataset of 694 instances.

## 3 Background

In this work, we evaluate various machine learning algorithms for the task of hypertension prediction. Specifically, we design neural networks with varying hyperparameters and structure and provide a comprehensive analysis of impact of these parameters on the performance. Further, we also compare to other machine learning methods to compare and contrast the effectiveness of various paradigms for such prediction. Prior to dwelling in the core designs, we provide a brief background of the techniques used.

### 3.1 Neural Networks

Neural Network is the machine learning algorithm that is designed to recognize patterns in the input data, learns from it and create a weight matrix on the basis of the relationship among the input parameters. Neural Network algorithm is inspired from the human brain and mimics the working of neurons. It has an input layer, some hidden layers and output layer. This architecture is shown in Fig 1. A neuron in human brains is replaced by a node in neural network, these nodes are interconnected with each other with some weights. These weights are computed by the algorithm itself during the training phase.

**Fig. 1:**
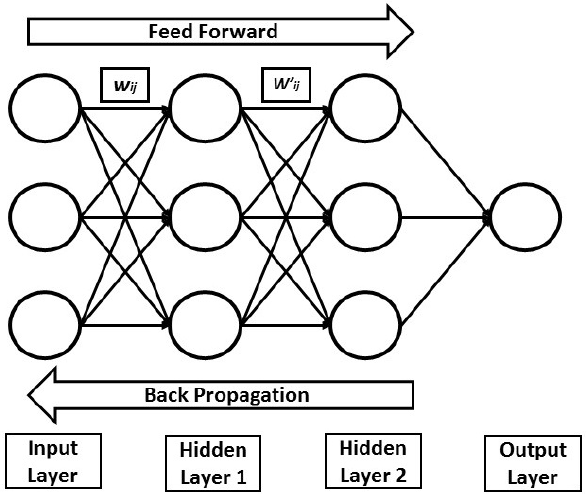
A simple Neural Network

The output in neural network is represented as the summation of product of all input values and their corresponding weights along with a bias passed through an activation function, as shown in the following equations:

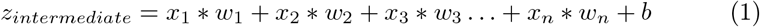

which can be written as,

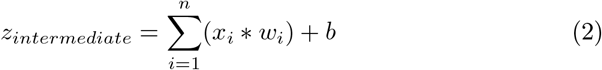

where *z*_*intermediate*_ is the output …, *n* is the size of input vector, *x*_*i*_ represents the *i*^*th*^ input, *w*_*i*_ is the weight of the *i*^*th*^ layer, *b* is the bias

Here bias is always 1 and has its own weight value. It is introduced so as to make sure that neuron is activated even when all the inputs are 0. This intermediate output is then passed through an activation function to introduce non linearity in the network. The model has two phases during the training, *viz*. forward propagation and backward propagation.

During the forward propagation, the inputs are provided in the input layer of the network and an output is acquired after passing it through hidden layer(s) and activation function. This output value is the predicted value. The forward propagation ends here and backward propagation starts. To verify the correctness of the predicted value we moves backwards in the network, updating the weights so as to minimize the error in the predicted value. The minimization of error is ensured by loss function. One forward propagation and one backward propagation is combinedly called an as epoch.

### REctified Linear Unit (relu)

It is the activation function which gives output as the maximum value between 0 and the input value. It discards the negative input value and changes it to 0. Mathematically,

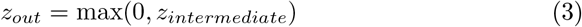

It is mathematically simple, therefore it is less resource intensive in implementation. Graphically it is represented in Figure 2.

**Fig. 2:**
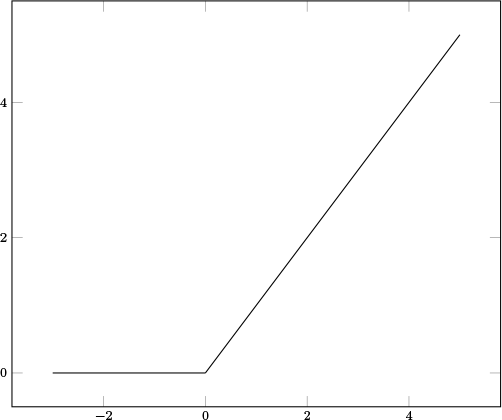
ReLU Activation Function

### Softmax

It is the activation function which is used in the output layer because of its ability to turn numbers into probabilities between 0 and 1. It is used in cases where more than one inputs are received and we need to check the maximum probability of occurrence among multiple inputs. Mathematically,

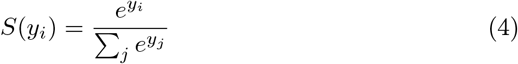

Where, *y*_*i*_ is the *i*^*th*^ input to the function. Graphically it is represented in Figure 3.

**Fig. 3:**
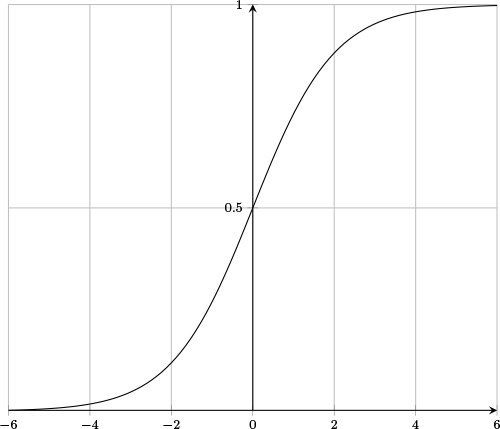
Softmax Activation Function

### 3.2 Other Algorithms

#### Support Vector Machine

Support Vector Machine is a supervised learning algorithm that classifies data on the basis of hyperplane that separates the dataset. During training the algorithm draws a hyperplane that linearly classifies the data into the number of classes. In case of binary classification, this plane is a line.

The SVM takes the labeled input as training data. This data is then used to create a plane that separates the data into labelled classes through the following process in case of binary classification:

- A linearly separable line is drawn between the classes.
- The nearest two points from the line are selected among both the classes. This is done by calculating distances among all the points and the line. They are called support vectors.
- The distance between support vectors and the plane is called margin. This margin is maximized.

The two hyperparameters in SVM are regularization and gamma. Regularization controls the smoothness of the plane. This translates as the margin of the hyperplane. More value of regularization tells the algorithm to learn every instance in the dataset, therefore smaller hyperplane. However high value of regularization also risks overfitting. Gamma defines the effect of the points that may cast influence over the hyperplane. Higher value of gamma the points closer to the hyperplane are taken into consideration wheres in low gamma value the points far from the hyperplane is taken into calculations.

#### Random Forest

Random forest is the classification algorithm that works on the principle of wisdom of crowd. The random forest relays on the fact that unrelated decision trees when combined produce better results. Decision trees are constructed as flowcharts and predict a final outcome. Random forest has two hyperparameters, Number of estimators and maximum depth. Number of estimators is the number of trees that the model is allowed to create. More the number of estimators more is the chance of higher accuracy but also is more computationally expensive and increases the risk of overfitting. Maximum depth represents the depth of each tree in the random forest. More the depth captures more data in the training instances. However it has to be taken into consideration the fact that higher depth also risks overfitting.

## 4 Designing a Neural Network

Neural Network contains multiple hyperparameters, such as number of layers, epochs number of neurons in each layer, activation function, etc. The balanced combination of all hyperparameters ensures a good neural network that identifies the patters in the data and predict the outcomes in the new data smoothly. Hyperparameter tuning is the most challenging task in any neural network design. Masanori Suganuma et. al. [15] have demonstrated the use of Cartesian genetic programming (CGP) in automatically designing a convolutional neural network. Dougal Maclaurin et. al. [16] uses the stochastic gradient descent to derive hyperparameters.

### How to choose layers in a neural network?

Less number of layers will underfit the model and more number of layers will overfit the model. Increasing the number of layers is also computationally expensive as more number of multiplication and addition will be required to get the output. However the number of layers and number of neurons varies greatly from problem to problem as explained by Imran Shafi et. al [17].

### What is the impact of neurons?

Number of neurons in the input layer is normally equal to the parameters in the training dataset. Neurons in hidden layers varies depending upon the learning required. Whereas output layer contains the number of neurons equal to the class variables.

### What is the impact of activation function?

Activation function is important in moulding the output in correct shape and induce non-linearity in the network. Without activation functions, the output of the neural network will be linear and thus a liner regression. There are multiple activation functions available such as Sigmoid Function, TanH Function, ReLU (Rectified Linear Unit), Leaky ReLU and Softmax. Evaluation of multiple activation functions is also done by researchers [18].

### Impact of Hyperparameters

Hyperparameter tuning is vital to get the meaningful predictions from the model. The graphs shows the relationship between accuracy and batch size, number of layers and number of neurons.

Figure 4(a) shows the impact of batch size on accuracy on open source dataset. The graphs shows initial increase with decrease in batch size which depicts the learning of the model. 128 batch size makes the model reaching an accuracy of more than 88%, while 16 batch size shows an accuracy of more than 91%. Upon reaching a saturation value, the accuracy flattens and if continued from here the risk of overfitting arises.

**Fig. 4:**
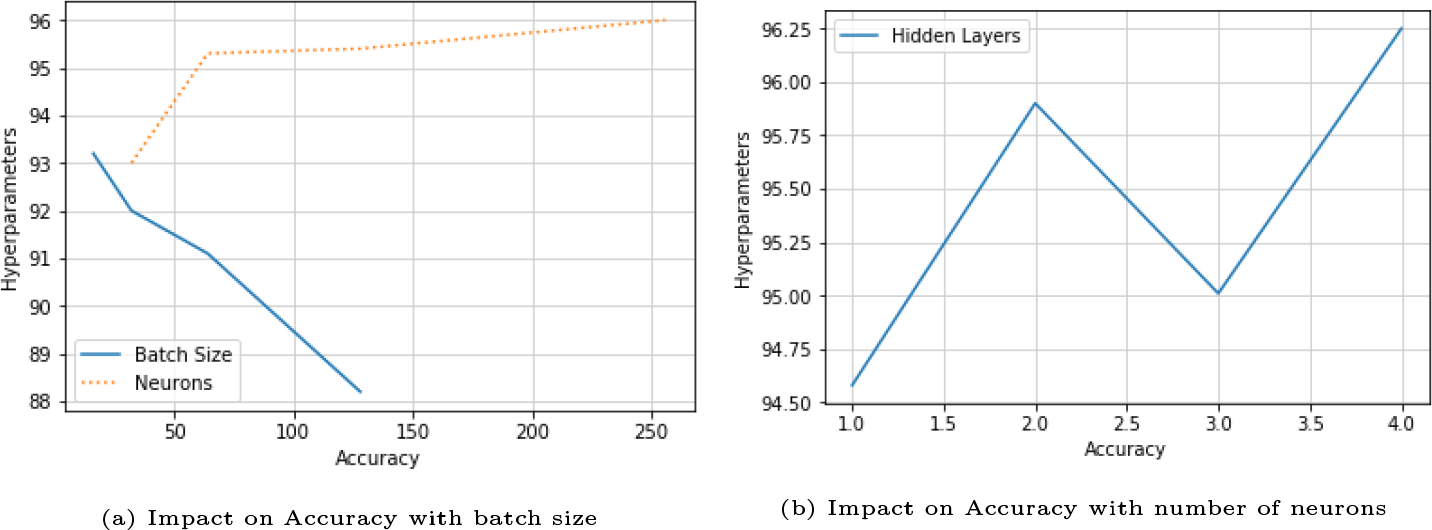
Impact of (a) batch size and number of neurons in the hidden layers and (b) number of layers on the performance of neural network with MNIST dataset.

Figure 4(a) shows the relationship between accuracy and number of neurons. More number of neurons increases the accuracy where 32 neurons has an accuracy of more than 93% and 256 neurons reaches an accuracy of more than 96%.

Figure 4(b) shows the impact of number of layers on accuracy. The graphs shows a decrease in accuracy with increase in number of layers with some fluctuations. However if this continues further, the accuracy flattens and condition of overfitting arises.

## 5 Experiments

### 5.1 Dataset

The data is collected from a tertiary care hospital in India. A set of questions were prepared and the data was collected from the patients. The dataset consists of 528 instances with 141 variables (such as age, religion, education, etc.). The dataset contains a mix of continuous and categorical values. Variables such as age, family income are continuous values and variables such as family type, housing type are categorical values. Much of the data is in numeric form except two variables.

Exploratory data analysis in the provided data shows the following trends with respect to age, alcohol consumption and average working hours:

### 5.2 Data Cleaning

- During the analysis of the data, dependent and derived parameters were removed.
- Parameters whose value remains unchanged were also dropped from the training data.
- The string value columns were label encoded.

After data cleaning 124 variables were left. The class variable is BPgroup which is categorical (0 or 1). The total positive instances are 134 and total negative instances are 394.

### 5.3 Implementation Details

#### Input Representation

The input vector consists of all the parameters along with their valid values. The problem is a binary classification problem where the output from the output layer will be 0 or 1, where 0 represents that patient is not suffering and 1 represents that patient is suffering from hypertension. The input vector is represented by,

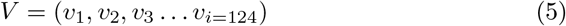

where *v*_*i*_ is the *i*^*th*^ parameter of the input vector. The total size of input vector is 124, hence the last parameter is *v*_*i*=124_.

#### Architectures

Table 3 shows the different machine learning algorithms that are evaluated *viz*. Neural Network, Support Vector Machine and Random Forest along with the parameters used for reporting the results in the paper. Further, we vary the number of hidden layers and neurons in the neural networks while discuss ablation studies on varying other parameters in Section 6.

**Table 1:** Summary of dataset for Hypertension analysis

|  | Number | Percentage |
| --- | --- | --- |
| Positive Instances | 134 | 25.38 |
| Negative Instances | 394 | 74.64 |
| Total Instances | 528 | 100 |

**Table 2:** Statistics of Age, Alcohol consumption and working hours in the Hypertension dataset

| AGE |  |  |
| --- | --- | --- |
|  | Age > 35 | Age < 35 |
| Positive Instances | 32.02% | 8.16% |
| Negative Instances | 67.97% | 91.83% |
| ALCOHOL |  |  |
|  | Yes | No |
| Positive Instances | 29.96% | 24.66% |
| Negative Instances | 70.73% | 75.33% |
| WORKING HOURS |  |  |
|  | WH > 8 | WH < 8 |
| Positive Instances | 23.13% | 26.14% |
| Negative Instances | 76.86% | 73.85% |

**Table 3:** Model parameters for various algorithms

|  |  |
| --- | --- |
| Neural Network 1 ($NN_1$) | Batch Size = 124 |
|  | Epochs = 45 |
|  | Layers = 2 with 50 neurons each |
|  | Activation Functions = relu |
| Neural Network 2 ($NN_2$) | Batch Size = 124 |
|  | Epochs = 45 |
|  | Layers = 3 with 50 neurons each |
|  | Activation Functions = relu |
| Neural Network 3 ($NN_3$) | Batch Size = 124 |
|  | Epochs = 45 |
|  | Layers = 4 with 50 neurons each |
|  | Activation Functions = relu |
| Neural Network 4 ($NN_4$) | Batch Size = 124 |
|  | Epochs = 45 |
|  | Layers = 5 with 50 neurons each |
|  | Activation Functions = relu |
| Support Vector Machine | Train Test Split Ratio = 60:40 |
| Random Forest 1 ($RF_1$) | Number of Estimator = 10 |
| Random Forest 2 ($RF_2$) | Number of Estimator 20 |
| Random Forest 3 ($RF_3$) | Number of Estimator = 30 |

**Table 4:** Confusion Matrix

|  | Predicted NO | Predicted YES |
| --- | --- | --- |
| Actual NO | TN | FP |
| Actual YES | FN | TP |

**Table 5:** Results on hypertension dataset using neural networks (*NN*_*i*_), SVM and Random Forests (*RF*_*i*_)

| Algorithm | Accuracy | Precision | Recall |
| --- | --- | --- | --- |
| $NN_1$ | 73.00 | 55.00 | 8.7 |
| $NN_2$ | **74.52** | **60.00** | **15.70** |
| $NN_3$ | 72.16 | 25.00 | 1.70 |
| $NN_4$ | 72.16 | 25.00 | 1.70 |
| SVM | 73.11 | NA | 0 |
| $RF_1$ | 69.33 | 38.00 | 24.00 |
| $RF_2$ | **69.81** | **40.54** | **26.31** |
| $RF_3$ | 68.86 | 37.83 | 24.56 |

### 5.4 Metrics

#### Accuracy

Accuracy is the fraction of the values predicted by the model that are correct.

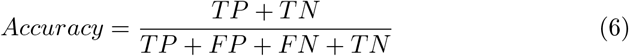

#### Precision

The fraction of the correct positive prediction against the overall predictions is called as Precision.

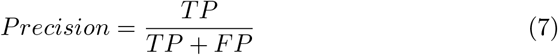

#### Recall (Sensitivity)

The fraction of correct positive prediction against overall observation in actual class is termed as Recall.

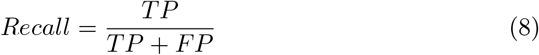

#### Confusion Matrix

Confusion matrix is a method of evaluation of the performance of a machine learning model. Confusion matrix is the table of predicted values against the actual values.

where, TP is True Positive, TN is True Negative, FP is False Positive and FN is False Negative.

## 6 Results

The neural network has maximum accuracy of 74.52% (Neural Network 2) followed by Random Forest at 69.81%. However, recall is maximum of Random Forest at 26.31. The graphs in Figure 5 shows the results where accuracy is plotted against epochs under varying hyperparameters. Figure 5 (a), (b) shows overfitting after 100 epochs. Figure 5 shows overfitting is a problem to tackle. For this, a regularisation technique dropout [19] is used in the model. Figure 5 (g) and (h) shows the impact of dropout.

**Fig. 5:**
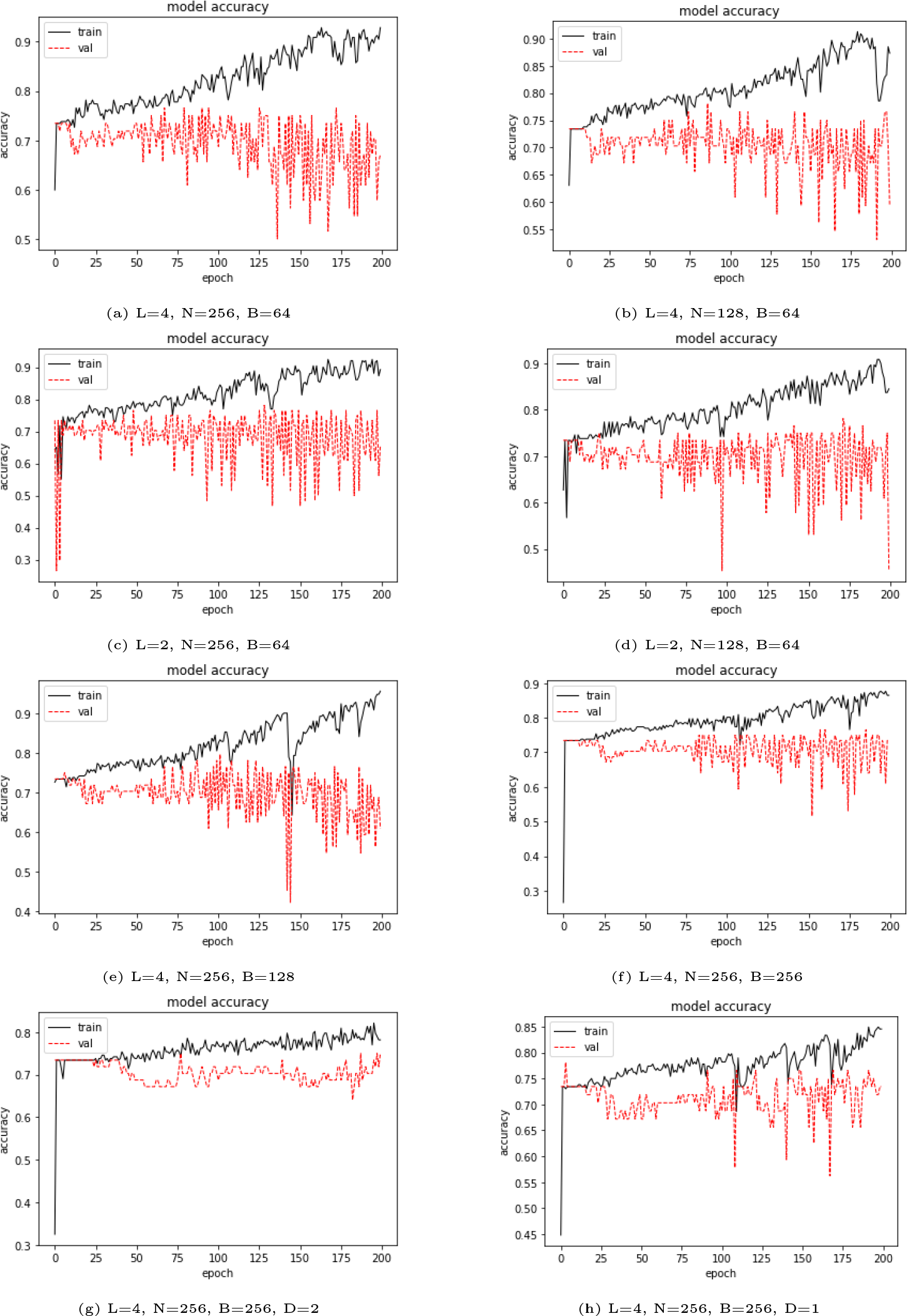
Ablation Study: Performance of the network on varying batch size, number of layers and number of neurons for predicting hypertension. Figure 5 (h) one dropout layer is included and the in Figure 5 (g) two dropout layers are included in the model.

As the results shows, neural network has outperformed both the algorithms. Neural Network 2, which has an accuracy of more than 75% is the best performing model. However recall is low in all cases that shows the weak sensitivity of the model. Increasing the recall remains the challenge in this dataset. Random forest also performed with more than 70% accuracy. However, random forest outperformed both the algorithms in terms of recall and precision. Random Forest 2 reaches a recall of more than 26.

## 7 Conclusion

We have studied the impact of various hyper-parameters on the performance of neural networks. The analysis was performed on MNIST dataset for analysing the impact while the findings have been used further on a custom dataset for hypertension prediction. We showed that while medical studies require large datasets, it is possible to provide reasonable performance with simple neural networks which can also generalize well.

## Data Availability

The data has been collected at the studied hospital

